# Public Awareness of Mask Usage in 29 Countries

**DOI:** 10.1101/2021.03.06.21253037

**Authors:** Susumu Annaka

## Abstract

This study analyzed the determinants of people wearing masks in 29 countries in 2020 using over 400,000 survey responses. The results showed that there is considerable variance in the awareness of mask usage across countries. People in Asian countries tend to wear masks whereas citizens in Scandinavian countries are less likely to wear them. This study also found that citizens in more democratic and more prosperous countries are less likely to wear masks. Furthermore, females are more likely to wear them. This paper contributes to the literature as it re-examines anecdotes and previous research by using statistical methods to analyze over 400,000 survey responses.

## 1. Introduction

The COVID-19 pandemic, which is the worst pandemic since the Spanish flu, has dramatically changed the world, with a significant number of people still suffering and dying from the disease. Without vaccination and magic bullet, one of the essential items that can protect people from the disease is a face mask^1 2^. The effectiveness of wearing masks has been reported in the United States^3^, Germany^4^, Hong Kong^5^, and other regions^6^. Yet, recognition of its importance varies across nations and even within a country. Some studies have also analyzed the characteristics that affect mask-wearing behavior, such as gender ^7^, age, the living place ^8^, and social norms ^9^. However, the studies of the effectiveness and determinants of wearing masks that were based on cross-national data were not thoroughly conducted, except for one^10^. Therefore, this study analyzed which people in which country have a greater awareness of wearing masks by utilizing the cross-national and multilevel data of over 400,000 persons.

To summarize the findings of this study, there is considerable variance in the awareness of mask usage across countries. People in more prosperous and more democratic countries are less likely to wear masks, even after controlling for positive cases, death cases, and other factors. It was also found that females were more likely to wear masks.

## 2. Data

Figure 1 shows the trends in the awareness of wearing face masks of over 400,000 people in 29 countries in 2020 using data from the Imperial College London YouGov Covid-19 Behaviour Tracker Data Hub ^11^. The graphs present each country’s monthly average response to the statement “Worn a face mask outside your home (e.g., when on public transport, going to a supermarket, going to a main road)”; the responses ranged from 1 (not at all) to 5 (always). The original order was reversed to aid understanding. The graphs in the figure reveal the following patterns: (1) there was a consistently high level of awareness of wearing masks throughout the year (Hong Kong, Indonesia, Malaysia, South Korea, and Thailand), (2) there was an increasing level of awareness (Canada, Denmark, Finland, the Netherlands, the U.K., and the U.S.), and (3) there were consistently low levels of awareness (Norway and Sweden). These patterns support previous research^10^.

**Figure 1.**
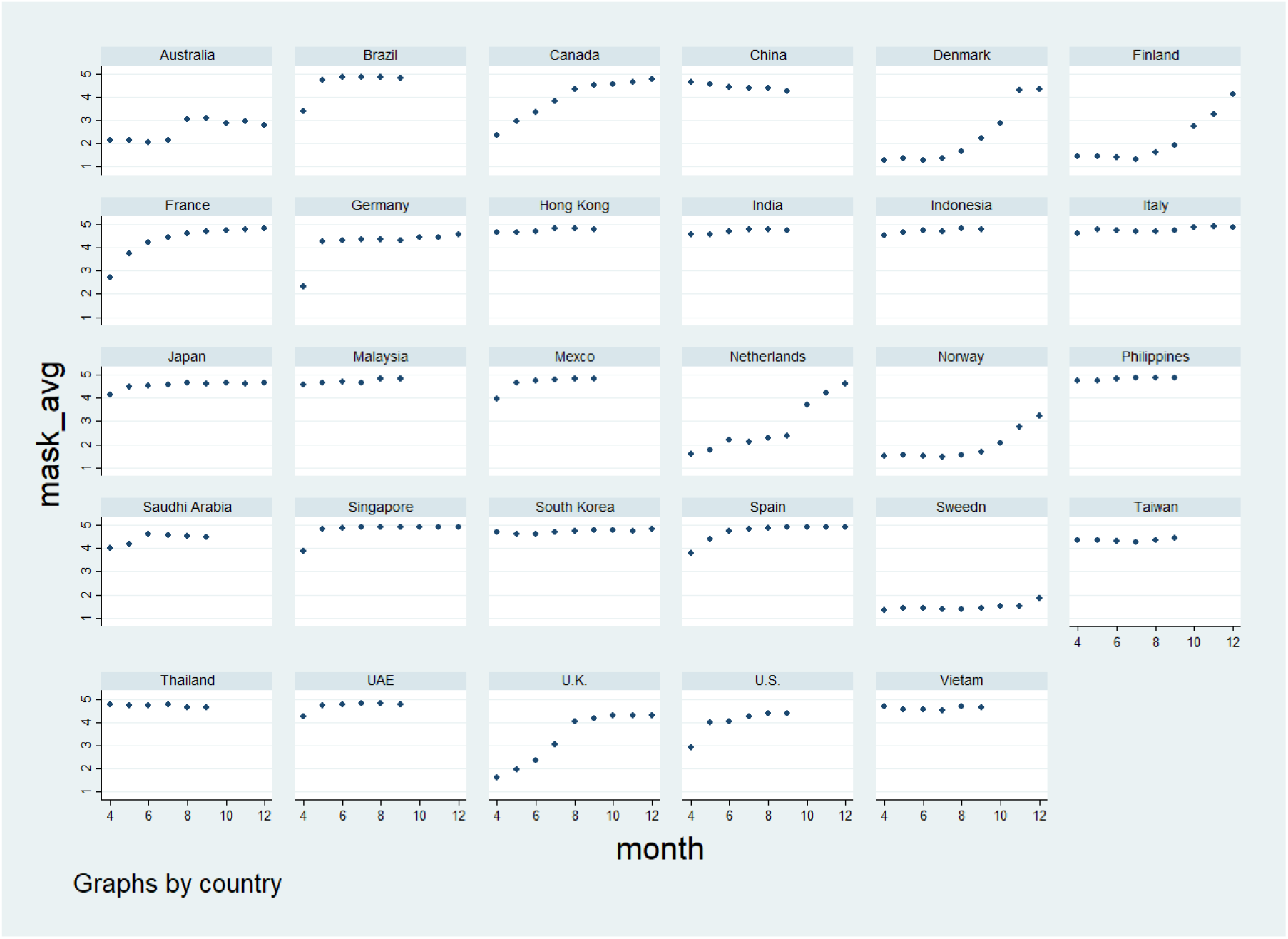
Trends in Wearing Masks by Country. Note: mask_avg is each country’s monthly average response to the statement “Worn a face mask outside your home (e.g., when on public transport, going to a supermarket, going to a main road)”; the responses ranged from 1 (not at all) to 5 (always). The original order was reversed to aid understanding.

It seems that people in Asian countries, in which relatively fewer COVID-19 cases and deaths have been reported, are more likely to wear masks in public spaces. On the other hand, people in Scandinavian countries, where individual freedom is regarded as especially important, are less likely to wear masks. Although the responses may have been affected by social desirability bias, it is certain that they present at least some precious knowledge on the issue. However, these trends did not consider the number of positive cases and deaths. In other words, people in less severely affected countries tend to wear masks. In the next section, we analyze the effects of these factors on the awareness of wearing masks.

## 3. Methods

### 3.1 Statistical model

In this section, the determinants of the awareness of wearing masks across countries are explored using statistical methods and considering other factors, such as the political regime, living standards, and population density at the national level. Two models were estimated separately to avoid multicollinearity. The equations were as follows:

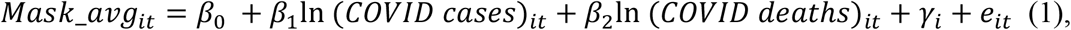

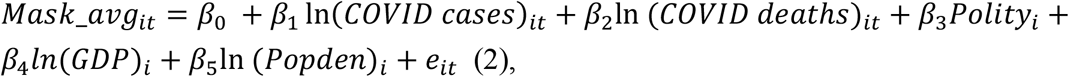

where, in Equation 1, *Mask*_*avg* is the country’s monthly average response to the statement “Worn a face mask outside your home (e.g., when on public transport, going to a supermarket, going to a main road)”; the responses ranged from 1 (not at all) to 5 (always). The original order was reversed. *COVID cases* and *deat*h*s* are the monthly averages of COVID-19 positive cases and death cases, respectively. These data were obtained from the Oxford COVID-19 Government Response Tracker^12^. *γ* represents the country-fixed effects, which are treated as independent variables. *e*_*it*_ is the error term of the regression, *i* is the country, and *t* is months. In Equation 2, *Polity* is the political regime variable that was taken from the Polity V project^13^. The Polity 2 variable was obtained from the project codes democracy levels that ranged from −10 (most autocratic) to 10 (most democratic). *GDP*, which is the gross domestic product (GDP) per capita, was used for living standards, and *Popden* is the population density, which were obtained from the World Development Indicators^14^. Except for Polity 2, the variables were logged. This equation does not include country dummy variables because their inclusion omits the time-invariant independent variables. Ordinary least squares was applied in both analyses. Appendix A1 presents the descriptive statistics.

Figure 2 shows the results of the county variation (the base category is the United Arab Emirates). These reveal the variation across countries after controlling for COVID-19 positive cases and death cases. Again, the analysis confirms the patterns suggested by Figure 1 (the table of the results of Model 1 is presented in Appendix A2).

**Figure 2.**
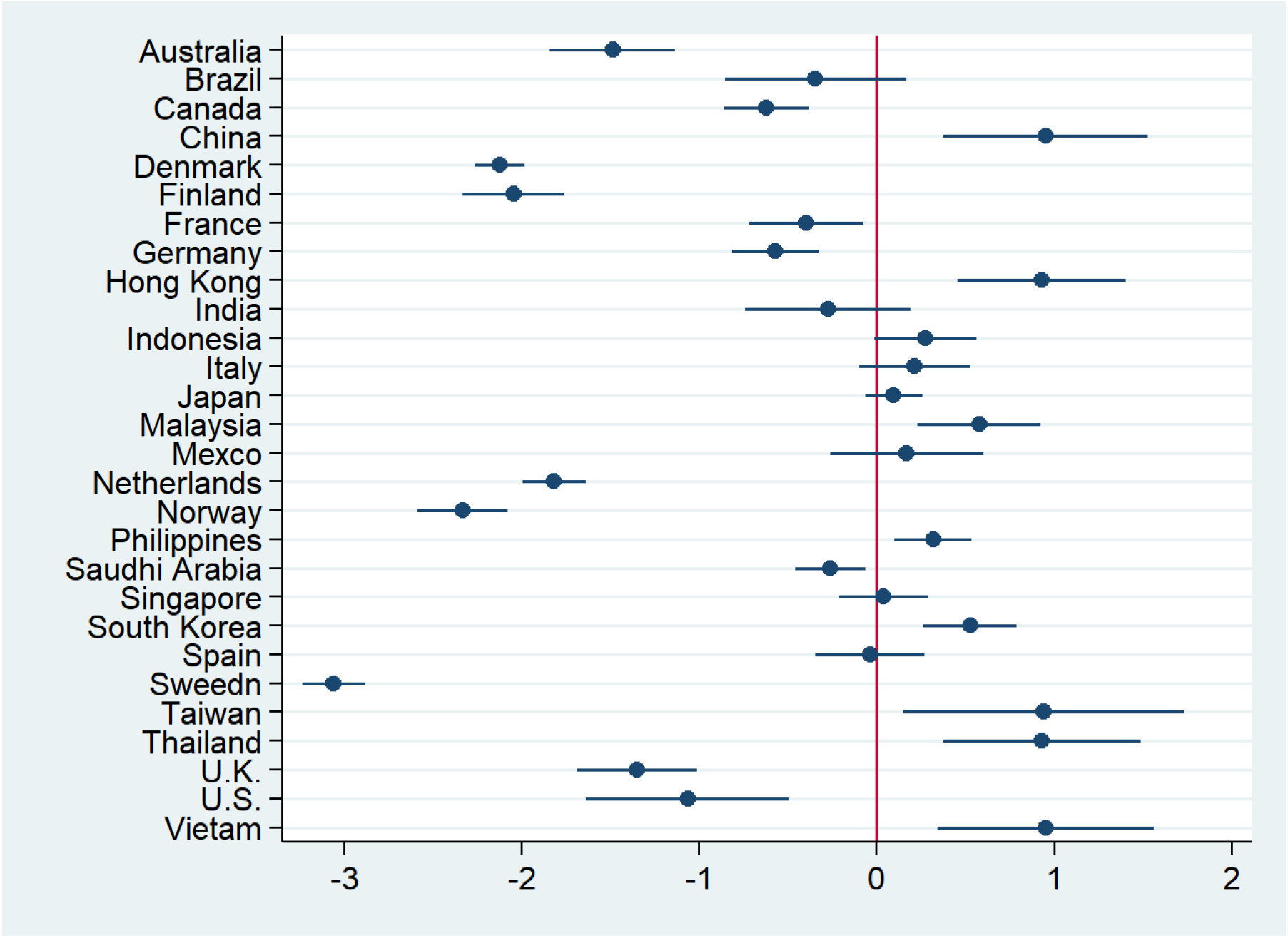
Results of the Regression of the Cross-National Differences in Wearing Masks. Note: 29 countries, *N* = 201, standard errors are clustered by country.

Figure 3 shows the results of the analysis including Polity2, GDP per capita, and population density. The table of the results of Model 2 is presented in Appendix A2. This figure indicates that there is a positive relationship between positive cases and mask usage, as expected. However, the relationship between the death cases and wearing masks was negative and not statistically significant. Polity 2 was negatively correlated with mask usage at a statistically significant level. This implies that people in democratic countries tend to wear masks less than those in authoritarian countries. GDP per capita was also negatively correlated with the awareness of wearing masks. On the other hand, population density was positively correlated, but was not statistically significant. These effects were expected. These results suggest that citizens in rich and democratic countries are less likely to wear masks.

**Figure 3.**
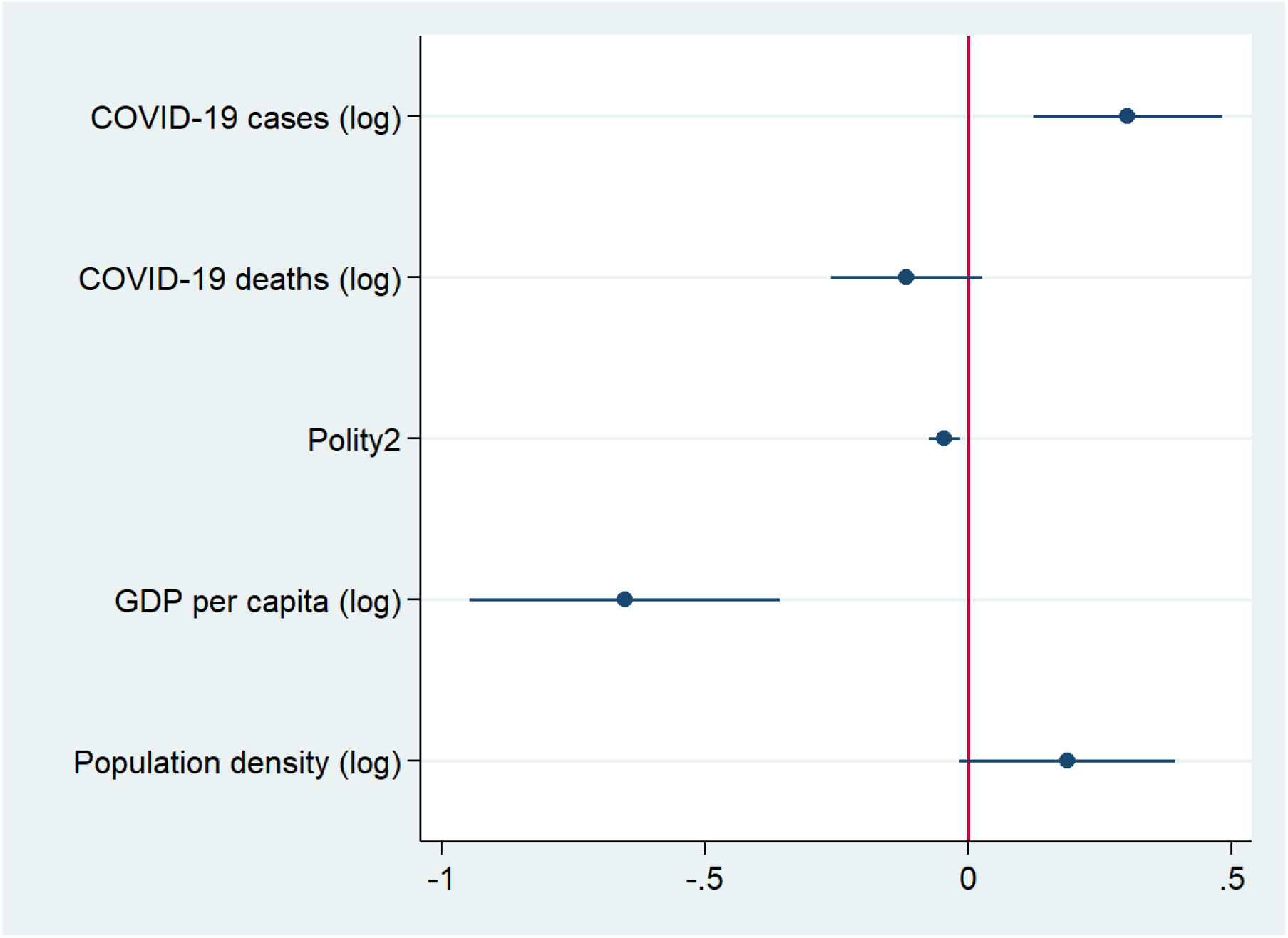
Results of the Regression of Other Cross-National Determinants of Wearing Masks. Note: 29 countries, *N* = 189, standard errors are clustered by country.

### 3.2 Multilevel analysis

In this section, we report a multilevel analysis based on a similar study^15^. Two models were estimated because of missing employment status values. However, the base model was the same. The equations were as follows:

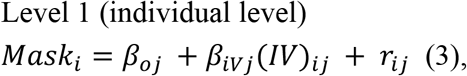

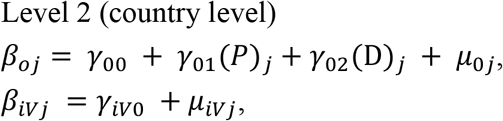

where *Mask*_*i*_ is the level of awareness of mask usage by person *i*, and *β*_*vj*_ is the intercept of the regression of country *j*. (*IV*)_*ij*_ is the vector of the independent variables (age, gender, and employment status), and *β*_*iVj*_is the regression coefficient of country *i. r*_*ij*_ is the error term of the regression. The intercept of the regression at the individual level is determined by the factors at the country level. Thus, at Level 2, *γ*_*00*_ is the intercept at the country level, and *γ*_*01*_ and *γ*_*02*_ are the coefficients of (*P*)_*j*_ *and* (D)_*j*_, which express the logged monthly average COVID-19 positive cases and deaths. *μ*_*0j*_ represents the error term of the regression. Because the regression coefficients at the individual level are also affected by the country level, the coefficients *β*_*iVj*_ of the independent variables also have intercepts *γ*_*iV0*_ and error terms *μ*_*iVj*_ at the country level.

Figures 4 and 5 show the results of the multilevel analyses, and the table of the results of Models 3 and 4 are presented in Appendix A3. Figures 4 indicates that COVID-19 positive cases and death cases had the opposite effect on the awareness of wearing masks, as shown in Figure 3. The coefficient of the confirmed cases had a positive sign, as expected, but that of the death cases was not positive. Age was not correlated with the awareness of mask usage. Males tended to wear masks less frequently than females. Figure 5 shows the results of the analysis that included the employment status. This variable was coded as 1 = full-time employment, 2 = part-time employment, 3 = full-time student, 4 = retired, 5 = unemployed, 6 = not working, and 7 = other. A higher number indicates isolation from society. This inclusion led to there being approximately 100,000 fewer samples for 7 countries due to missing values for the employment question. The coefficient of the variable suggests that people who are isolated from society are less likely to wear masks.

**Figure 4.**
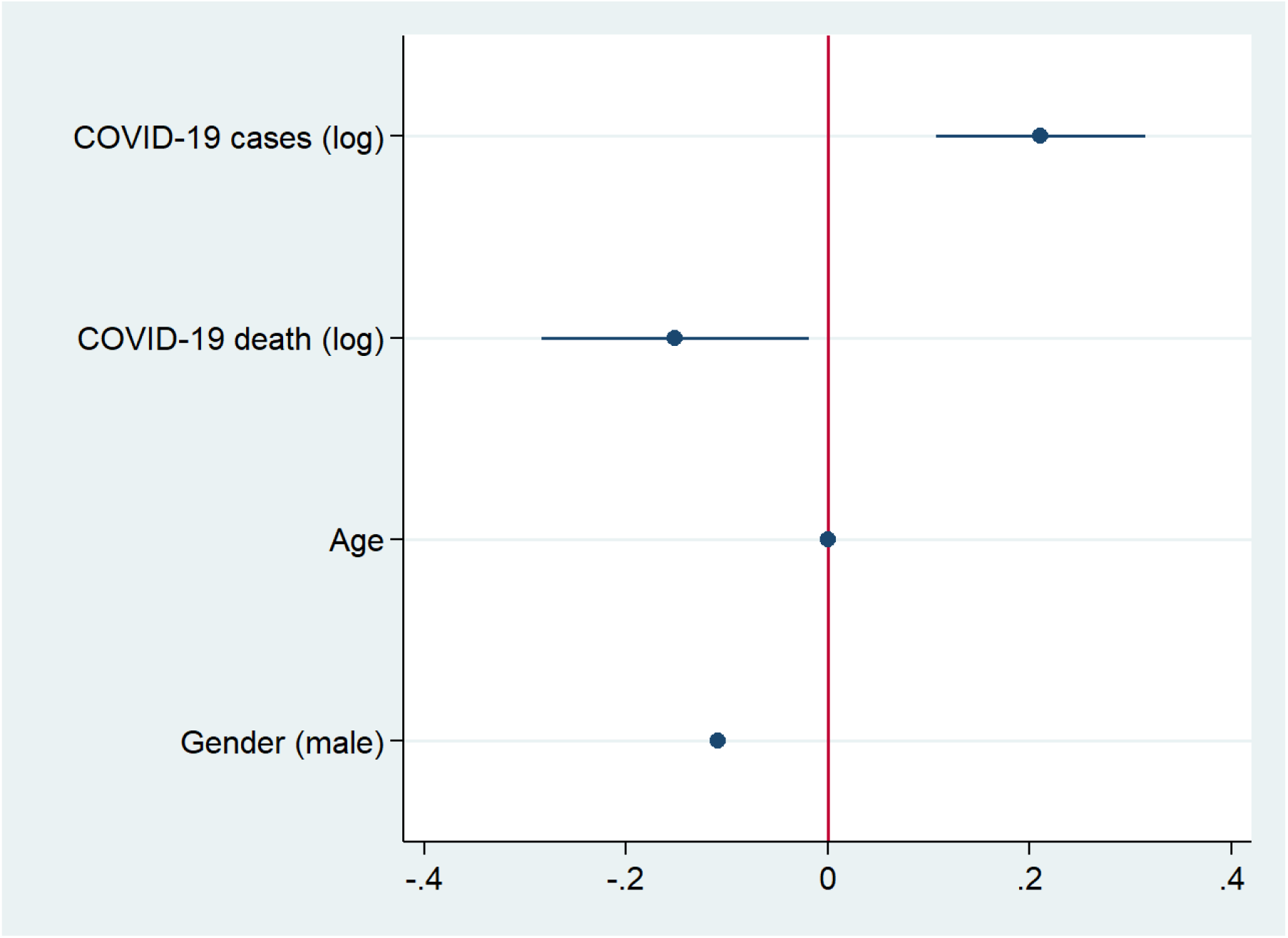
Multilevel Results of Other Cross-National Determinants of Wearing Masks. Note: 29 countries, *N* = 413,042.

**Figure 5.**
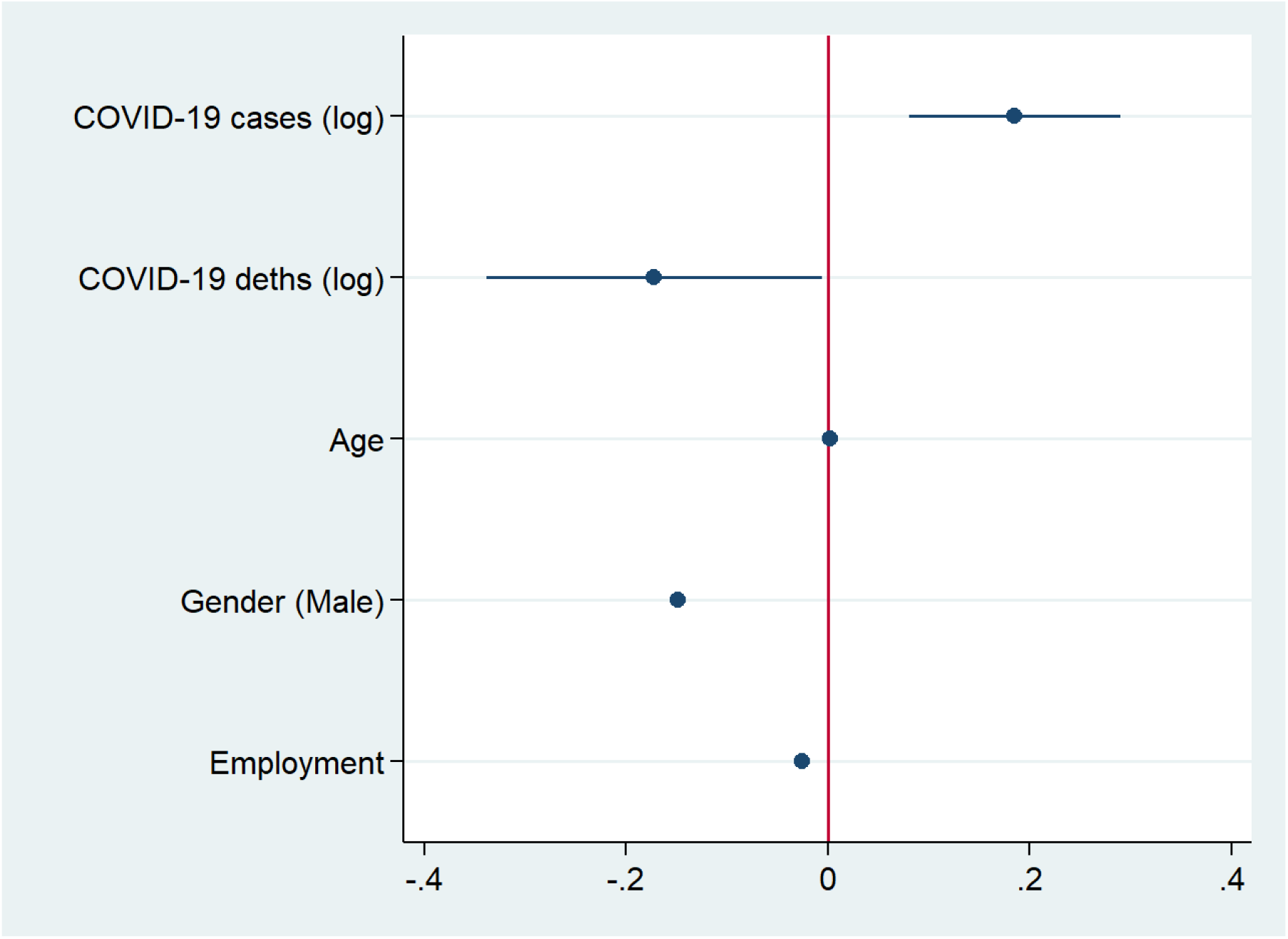
Multilevel Results of Other Cross-National Determinants of Wearing Masks Including Employment. Note: 22 countries, *N* = 322,749.

## 4. Discussion

This study found clearly different patterns of mask usage awareness, and the data presented in Figures 1 and 2 support a previous cross-national survey analysis^10^. People in Asian countries tend to wear masks whereas citizens in Scandinavian countries are less likely to wear them. This article also reports that citizens in more democratic and more prosperous countries are less likely to wear masks. While this result is anecdotally known, this study confirms it using a statistical method. This study also revealed that confirmed COVID-19 cases and death cases have the opposite effect on mask usage awareness. This finding may imply that people are cautious about the virus when there is a growing number of positive cases. On the other hand, the variable of the death cases may include some time lags. However, a previous study in Spain reported a null result for the effect of positive cases^9^. The research shows a much stronger effect of the average awareness of using masks at both the regional and provincial levels than that of positive cases. Figures 3 and 4 also support the gender effect, which has been found in a previous research study^7^, although another study did not confirm this^9^. The figures do not indicate that there is a consistent effect of age on wearing masks, which previous studies have also reported^8 9^. Figure 4 shows the effect of the employment status.

## 5. Conclusion

This study analyzed the determinants of wearing masks in 29 countries in 2020 using over 400,000 survey responses. The results show that there is considerable variance in the awareness of mask usage across the countries. While people in Asian countries tend to wear masks, citizens in Scandinavian countries are less likely to wear them. This article also reports that citizens in more democratic and more prosperous countries are less likely to wear masks. In addition, females are more likely to wear masks. This paper contributes to the literature as it re-examines anecdotes and previous research by using statistical methods to analyze over 400,000 survey responses.

## Data Availability

All the data used are available online.

---

**Appendix A1.** Descriptive statistics.

| Variable | Obs | Mean | Std. Dev. | Min | Max |
| --- | --- | --- | --- | --- | --- |
| Mask usage average per month | 219 | 3.2056 | 0.3308 | 2.5592 | 3.7212 |
| Average confirmed cases per month(log) | 219 | 5.9500 | 2.7957 | -1.7918 | 11.3780 |
| Average confirmed deaths per month(log) | 201 | 2.2912 | 2.9249 | -3.4340 | 7.6133 |
| Polity2 | 213 | 6.9155 | 5.6043 | -10 | 10 |
| GDP per capita (log) | 207 | 10.2826 | 0.9535 | 7.5830 | 11.4308 |
| Population density (log) | 213 | 4.7550 | 1.8299 | 1.1780 | 8.9813 |
| Multilevel |  |  |  |  |  |
| Mask usage | 448482 | 3.8072 | 1.6251 | 1 | 5 |
| Average confirmed cases per month(log) | 448541 | 6.0395 | 2.7691 | -1.7918 | 11.3780 |
| Average confirmed deaths per month(log) | 413101 | 2.5381 | 2.9875 | -3.4340 | 7.6133 |
| Age | 448541 | 43.5066 | 16.3714 | 18 | 99 |
| Gender | 448541 | 0.4988 | 0.5000 | 0 | 1 |
| Employment status | 335197 | 2.6609 | 1.8867 | 1 | 7 |

**Appendix A2.** Regression results for Figures 2 and 3.

| VARIABLES | (1)<br>Mask avg | (2)<br>Mask avg |
| --- | --- | --- |
| COVID-19 cases (log) | 0.319***<br>(0.0847) | 0.303***<br>(0.0869) |
| COVID-19 deaths (log) | -0.159***<br>(0.0553) | -0.117<br>(0.0698) |
| Polity2 |  | -0.0444***<br>(0.0146) |
| GDP per capita (log) |  | -0.652***<br>(0.143) |
| Population density (log) |  | 0.189*<br>(0.0998) |
| Countries | Y | N |
| Constant | 2.812***<br>(0.500) | 8.354***<br>(1.335) |
| Observations | 201 | 189 |
| R-squared | 0.842 | 0.501 |

Robust standard errors in parentheses
\*\*\* p<0.01, \*\* p<0.05, \* p<0.1

**Appendix A3.** Regression results for Figures 4 and 5.

| VARIABLES | (3)<br>Mask | (4)<br>Mask |
| --- | --- | --- |
| COVID-19 cases (log) | 0.211***<br>(0.0532) | 0.185***<br>(0.0532) |
| COVID-19 deaths (log) | -0.151**<br>(0.0677) | -0.172**<br>(0.0848) |
| Age | -0.000127<br>(0.000115) | 0.00211***<br>(0.000134) |
| Gender (male) | -0.109***<br>(0.00354) | -0.149***<br>(0.00406) |
| Employment status |  | -0.0254***<br>(0.00110) |
| Constant | 3.112***<br>(0.501) | 3.569***<br>(0.542) |
| Observations | 413,042 | 322,749 |
| Number of groups | 29 | 22 |
| Standard errors in parentheses |  |  |
| *** p<0.01, ** p<0.05, * p<0.1 |  |  |

